# OQA : A question-answering dataset on orthodontic literature

**DOI:** 10.1101/2024.07.05.24309412

**Authors:** Maxime Rousseau, Amal Zouaq, Nelly Huynh

## Abstract

**Background:** The near-exponential increase in the number of publications in orthodontics poses a challenge for efficient literature appraisal and evidence-based practice. Language models (LM) have the potential, through their question-answering fine-tuning, to assist clinicians and researchers in critical appraisal of scientific information and thus to improve decision-making.

**Methods:** This paper introduces OrthodonticQA (OQA), the first question-answering dataset in the field of dentistry which is made publicly available under a permissive license. A framework is proposed which includes utilization of PICO information and templates for question formulation, demonstrating their broader applicability across various specialties within dentistry and healthcare. A selection of transformer LMs were trained on OQA to set performance baselines.

**Results:** The best model achieved a mean F1 score of 77.61 (SD 0.26) and a score of 100/114 (87.72%) on human evaluation. Furthermore, when exploring performance according to grouped subtopics within the field of orthodontics, it was found that for all LMs the performance can vary considerably across topics.

**Conclusion:** Our findings highlight the importance of subtopic evaluation and superior performance of paired domain specific model and tokenizer.

## 1 Introduction

In the contemporary era, the adoption of evidence-based practice has emerged as the prevailing paradigm guiding the provision of optimal therapeutic interventions ubiquitously across diverse domains of healthcare, including dentistry and its various specialties [1–3]. According to this principle, clinicians ought to ground their decisions on scientific knowledge coming from peer-reviewed publications and take into consideration study design and statistical rigor [4]. In recent years, the number of scientific publications has increased exponentially. Systematic reviews and meta-analyses cannot keep up with the current pace of novel research publications [5]. This, in turn, leads to information overload for clinicians [6].

Recent advances in artificial intelligence (AI) have shown promising results of language models (LMs) on various natural language processing (NLP) tasks in the biomedical domain, including question-answering (QA) [7]. If we consider peer-reviewed literature as an unstructured knowledge base, LM could be used to assist clinicians by querying the literature in the form of extractive QA.

This paper aims to investigate current LMs’ capabilities for such a task in the domain of orthodontics. To this end, the first expert-curated QA dataset in the field of orthodontics, along with an assessment of the capabilities of various LMs. Insights on the training and evaluation of language models are provided. Although this study focused on the topic of orthodontics, the findings outlined in this paper should prove useful to all fields of dentistry. In addition, the dataset and code are made available to bootstrap research efforts in the oral health community.

## 2 Related-work

The introduction of the Transformer architecture [8] has led to considerable advances in general domain NLP that have been adapted to the biomedical field. When comparing models of similar parameter counts (i.e. model size), biomedical models tend to outperform general domain transformers on named entity recognition (NER), relation extraction, document classification, sentence similarity, and QA [7]. Interestingly, it seems that even large language models (LLMs) such as GPT-3, which demonstrates impressive zero-shot capabilities, also suffer from performance issues in the biomedical domain [9] and still require in-domain fine-tuning to achieve strong performance [10].

Various studies have used general domain pretrained transformers and performed further pretraining on a biomedical corpus before fine-tuning on a downstream biomedical task. This is known as continual pretraining [11]. However, some authors [7] claim that pretraining from scratch is superior to continual pretraining.

There are four major categories of QA datasets in the biomedical field: examination, clinical, scientific, and public information [12]. Examination datasets such as HEAD-QA [13] are used to test a system’s knowledge and are usually created from question banks meant to assess students. These datasets generally require retriever models to gather useful information and provide multiple choices for answers. Clinical datasets such as CliCR [14] are used to assess patient history of case reports to answer a given query. Datasets such as these aim to assist clinical decision-making. Public information datasets like MEDIQA-AnS [15] are concerned with answering questions for laypersons. Scientific QA datasets, such as BioASQ [16], aim to extract answers from scientific literature and test for machine reading comprehension capabilities. The safe deployment of LMs in real-world settings hinges on exhaustive evaluations. Existing biomedical datasets do not contain samples related to dentistry. Therefore, there is a need to create datasets specific to the field of dentistry to assess model performance.

Most large-scale datasets crowdsource their data curation process, such as the Stanford Question Answering Dataset (SQuAD) [19]. However, this is very difficult to do when domain expertise is needed. Although some large biomedical multiple-choice and true/false QA datasets exist [12], extractive scientific QA datasets only contain a few examples (Table 1). This is due to the greater amount of human labor required for their curation.

**Table 1.**
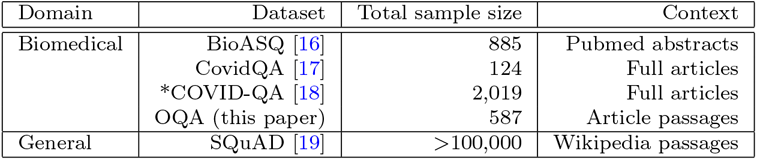
Comparison of available general domain and biomedical scientific question-answering datasets. *Open-Domain: this task included a document retrieval component.

| Domain | Dataset | Total sample size | Context |
| --- | --- | --- | --- |
| Biomedical | BioASQ [16] | 885 | Pubmed abstracts |
|  | CovidQA [17] | 124 | Full articles |
|  | *COVID-QA [18] | 2,019 | Full articles |
|  | OQA (this paper) | 587 | Article passages |
| General | SQuAD [19] | >100,000 | Wikipedia passages |

## 3 Methods

This study focused on the closed-book extractive QA task. This entails that, for each sample, the model receives the question and a passage as inputs and must extract the answer from the passage. Given limited resources for data curation, the dataset presented in this paper is composed of samples that pertain to the orthodontic specialty. It is well suited to gauge current LM capabilities as a variety of topics are covered (i.e., biomechanics, growth and development, biology of tooth movement, etc.). This dataset was created in a semi-automated fashion, with a human expert validating each sample. The experiments aim to set performance baselines for the proposed dataset. Both generative (sequence-to-sequence) and predictive (encoder-only) architectures were included. These two types of models differ in their outputs. Encoder-only models, such as BERT, output a probability logit for each token to correspond to the start and end position of the answer. Generative models will output a sequence of tokens from their vocabulary as their answer.

### 3.1 Dataset creation

OrthodonticQA (OQA) is a scientific extractive question-answering dataset specific to the field of orthodontics. The dataset is made openly available under the Apache-2.0 license ^1^. OQA was built with a format that is akin to the Stanford Question-Answering Dataset (SQuAD). SQuAD is a large-scale question-answering dataset that was created using crowdsourced curators. A sample is composed of a context, a question, and an answer. The context constitutes a passage from the corpus. The answer to the question is a span of text extracted from the context. The length of the context was limited to 512 space-delimited tokens, as most LMs currently have limited input sequence lengths. The questions, answers, and contexts from OQA are longer than those of the SQuaD dataset. Table 2 provides a summary of the composition of OQA.

**Table 2.** Summary of dataset composition. † : Number of samples. ‡ : mean space-delimited tokens and standard deviation.

|  |  |  |
| --- | --- | --- |
| †Topics | Clinical | 245 |
|  | Growth | 83 |
|  | Pathology | 78 |
|  | Materials | 64 |
|  | Anatomy | 45 |
|  | Biomechanics | 38 |
|  | Biology | 31 |
|  | Other | 3 |
| †Type | Relationship | 118 |
|  | Factual | 189 |
|  | Explanatory | 280 |
| †PICO | Included | 172 |
|  | Not applicable | 415 |
| ‡Length (mean, SD) | Question | 17.20 (5.49) |
|  | Context | 371.14 (103.54) |
|  | Answer | 12.98 (11.28) |

A semi-automatic approach was used to generate OQA. This process involved four steps (Figure 1).

**Fig. 1.**
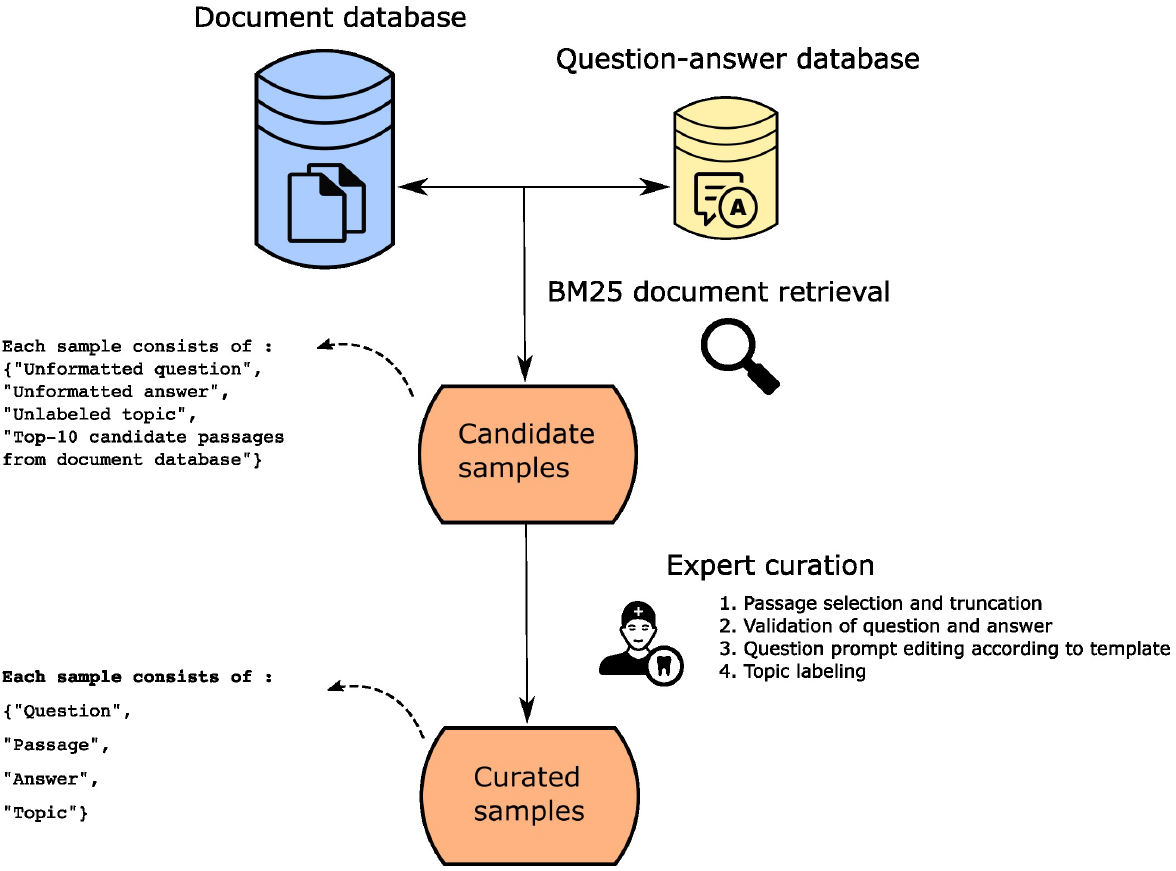
Overview of the dataset curation process

First, a corpus of over 6204 open-access orthodontic articles was collected. The included papers were published in The Angle Orthodontist ^2^ between January 1931 and January 2022. An index of the collection is released with the dataset for reproducibility purposes. Although there are more open-access publications in orthodontics, a smaller corpus allowed for more efficient document ranking on the available hardware for this study. Second, 2166 candidate questions were collected from online quiz platforms such as Anki, cram, and study-stack. Only sets of questions that pertained to the topic of orthodontics, either at an undergraduate or graduate level, were included.

Third, for each question, the top 10 candidate passages were extracted from the corpus using BM25 [20]. BM25 is a probabilistic ranking algorithm used for assessing the relevance of documents to a given query. It operates based on the similarity between the query terms and the terms present in the documents. This similarity is calculated while considering both the term frequencies in the documents and the information saturation of the terms in the entire corpus. Finally, a human investigator (MR) with domain expertise curated the samples. During the curation process, the passages were truncated to meet the context length limit. The questions and answers were validated and edited. Each sample was categorized according to a topic: materials, biomechanics, anatomy, biology, growth, clinical, and pathology (Figure 2). These were used to balance our training, validation, and test splits and assess performance on each topic. To validate the quality of our samples, an experienced orthodontist assessed a randomly chosen subset, which constituted 10% of the entire dataset. 93% were considered very clear, 7% were moderately clear and none were unclear.

**Fig. 2.**
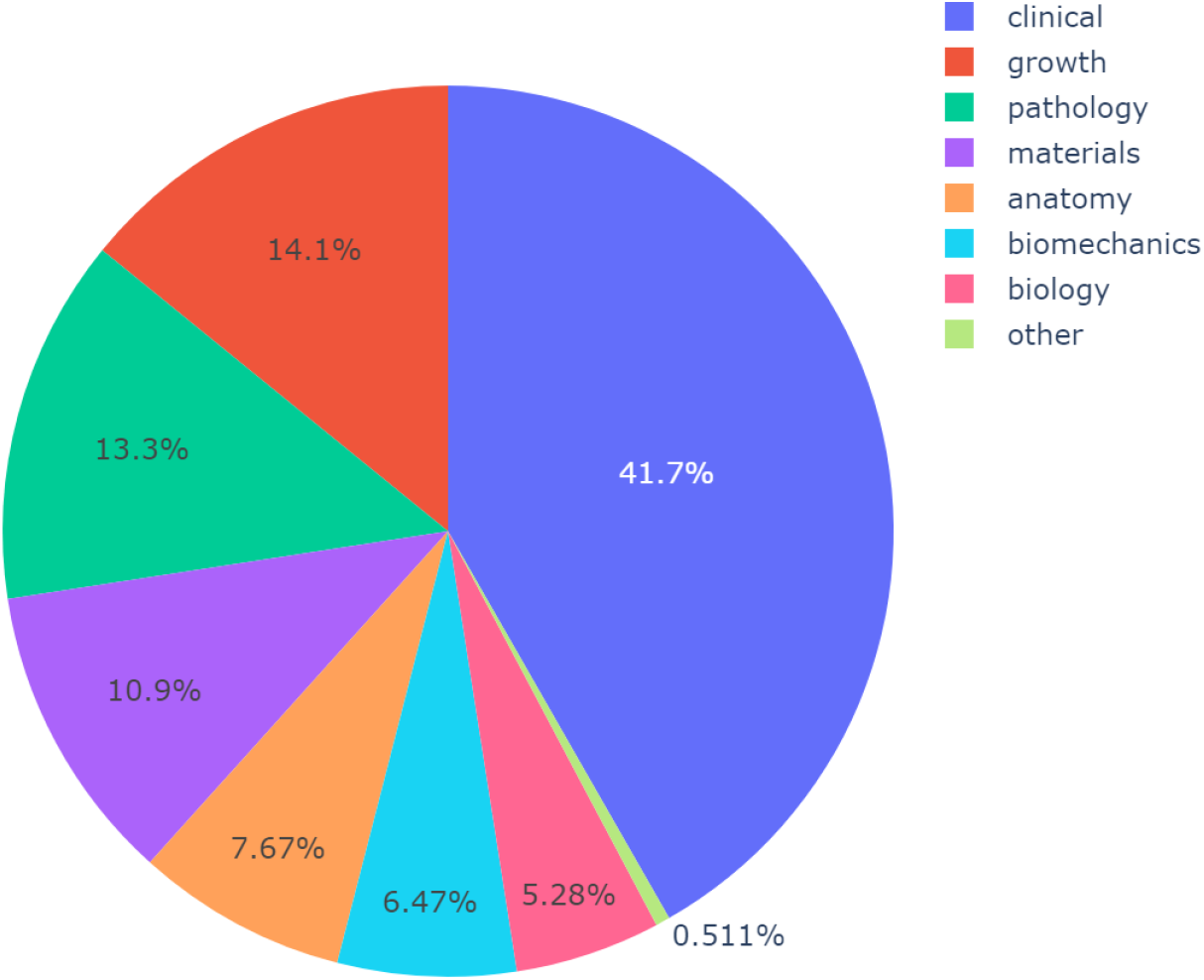
Distribution of topics from the full OQA dataset (including train, val and test sets.

We designed a style guide for the formulation of our samples to increase uniformity within our dataset (Table 3). Each question has two components: a preamble and a question statement. The preamble was formulated according to the topic of the sample to provide sufficient context to answer precisely. The PICO [21] elements were included in the preamble and/or question when possible. The question statement was formulated following its type (comparison/relationship, factual or explanatory). We consider that this standardized approach can be applied beyond the field of orthodontics.

**Table 3.**
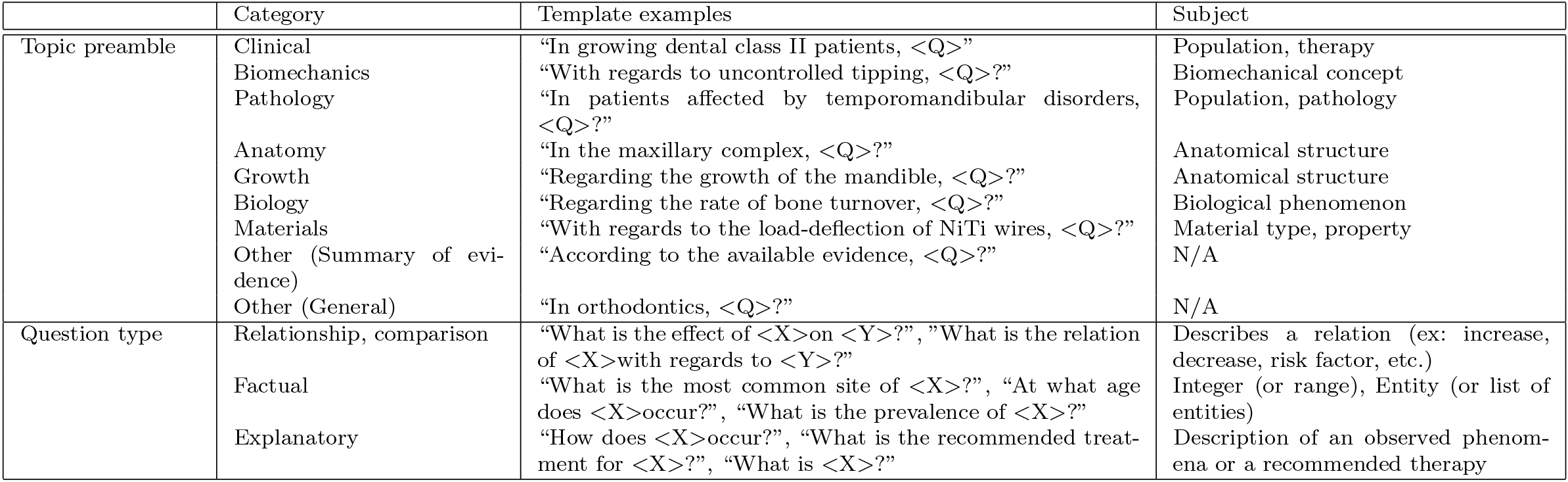
Overview of the preamble and question style guides used for the creation of the OQA dataset.

It is important to note that the dataset should be viewed as a reading comprehension assessment. This entails that the answer for a given sample corresponds only to that which can be extracted from the context, and may not reflect the current gold standard found in a systematic review on the topic.

### 3.2 Experimental setup

The OQA dataset was separated into three subsets for training, validation, and testing which contain 381, 92, and 114 samples respectively. Each subset contains a similar proportion of samples per topic. The validation set was used for hyperparameter tuning. After training, inference was performed on the test set to evaluate the performance of each model. The token-level F1 score and exact match were used to assess the model performances. The exact match corresponds to the percentage of samples where the model output is identical to the expected gold answer. The F1-score is a common metric in assessing QA performance [19], it is computed as the harmonic mean of precision and recall at the individual token level (Figure 3). Human evaluation of outputs was also conducted to qualitatively assess the best model.

**Fig. 3.**
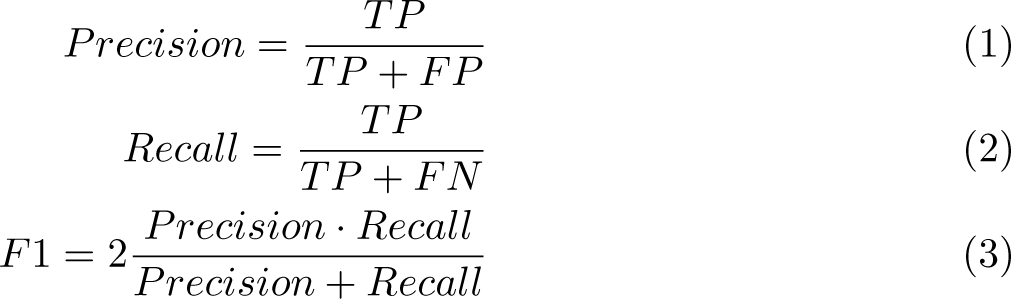
Precision, Recall and F1-score formulae. (TP) true positive, number of tokens predicted is present in the answer. (FP) false positive, number of tokens predicted that are not in the answer. (FN) false negative, number of tokens present in the answer that were not predicted.

A diverse selection of models was included for fine-tuning. Three encoder models: BERT [22], BioBERT [11], and PubmedBERT [7] were included. These models have different pretraining approaches: general domain, continual domain-specific pretraining, and domain-specific pretraining from scratch respectively. This allowed us to compare the influence of the type of pretraining. The BART [23] and T5 [24] generative models were also selected. This was motivated by the potential gains reported in the literature from the alignment of the pretraining objectives and downstream objectives on small datasets [25]. It is important to note however that T5 differs from BART as it was also trained on a large dataset containing diverse downstream tasks including QA. Additionally, two biomedical variants of BART and T5, BioBART [26] and SciFive [27] respectively, were incorporated into the experiments. These models were continually pretrained on a biomedical corpus. We fine-tune all generative models using the prompting technique proposed by Chada and Natarajan [25]. Finally, following the recommendation on biomedical QA model tuning [28], the best BERT-like model was pre-finetuned on SQuAD. This allows the investigation of the potential benefits of large-scale general-domain datasets for tasks related to dentistry. The code is released under the open-source Apache-2.0^3^. The mean and standard deviation for each model are reported in the result section.

All fine-tuning runs were performed with 12 epochs, a batch size of 8 for BERT-like models and 4 for T5 and BART models, and a learning rate of 2e-5 for all models except for the T5 and SciFive models where 1e-4 was used. Linear weight decay with a warm-up set to 10% of the total number of steps was used with the AdamW optimizer. The training checkpoint which attained the highest F1 score on the validation set during training was used for evaluation. The experiments were conducted using Python 3.10 and the PyTorch deep learning framework. The models were trained on a single NVIDIA V100 GPU with 16GB of memory. For each experiment, fine-tuning was performed with three different seeds (0, 10, 100).

## 4 Results

The performance of the various models fine-tuned on OQA is summarized in Table 4. The best model was PubmedBERT_SQuAD_, which achieved an F1 score of 77.61%. A considerable gain in performance was achieved by pre-finetuning the Pubmed-BERT model on a larger general domain QA dataset (SQuAD). T5, which was trained on a collection of tasks, also showed good performance with an F1 score compared to models trained only with the mask-filling objective (BART, BioBART, and SciFive). Domain-specific pretraining and vocabulary lead to increased performance for BioBERT, PubmedBERT, and BioBART. Post-hoc analyses illustrate large variations in performance based on the topic of the question for all models (Figure 4). PubmedBERT_SQuAD_, performs most evenly across topics of the test set. Biology, biomechanics, and pathology were amongst the topics where performance was generally lower.

**Table 4.** Performance of language models on the OrthodonticQA dataset. EM: exact-match, F1: F1-score, Size: model size by number of trainable parameters (millions). † : Biomedical domain vocabulary. ‡ : Generative model.

| Model | Size | Pretraining corpus | Fine-tuning datasets | EM mean (SD) | F1 mean (SD) |
| --- | --- | --- | --- | --- | --- |
| †PubmedBERT<sub>SQuAD</sub> | 110M | PMC (90GB) + Pubmed abstracts (21GB) | SQuAD | 53.22 (1.34) | 77.61 (0.26) |
| ‡T5 | 220M | C4 (745GB) | CoLA, SST2, MRPC, STS-B, QQP, MNLI, QNLI, RTE, CB, COPA, WIC, MultiRC, ReCoRD, BoolQ | 49.71 (2.68) | 72.75 (1.15) |
| ‡SciFive | 220M | C4 (745GB) + PMC (90GB) + Pubmed abstracts (21GB) | N/A | 38.89 (2.21) | 65.80 (1.62) |
| †PubmedBERT | 110M | PMC (90GB) + Pubmed abstracts (21GB) | N/A | 38.60 (2.32) | 61.54 (2.98) |
| ‡BioBART | 139M | RoBERTa (160GB) + Pubmed abstracts (21GB) | N/A | 26.90 (4.50) | 57.77 (3.23) |
| ‡BART | 139M | RoBERTa(160GB) | N/A | 29.53 (2.68) | 57.38 (1.24) |
| BioBERT | 110M | Wikipedia/TBC (20GB) + Pubmed abstracts (21GB) | N/A | 24.27 (4.99) | 48.30 (2.93) |
| BERT | 110M | Wikipedia/TBC (20GB) | N/A | 14.33 (3.32) | 37.13 (2.23) |

**Fig. 4.**
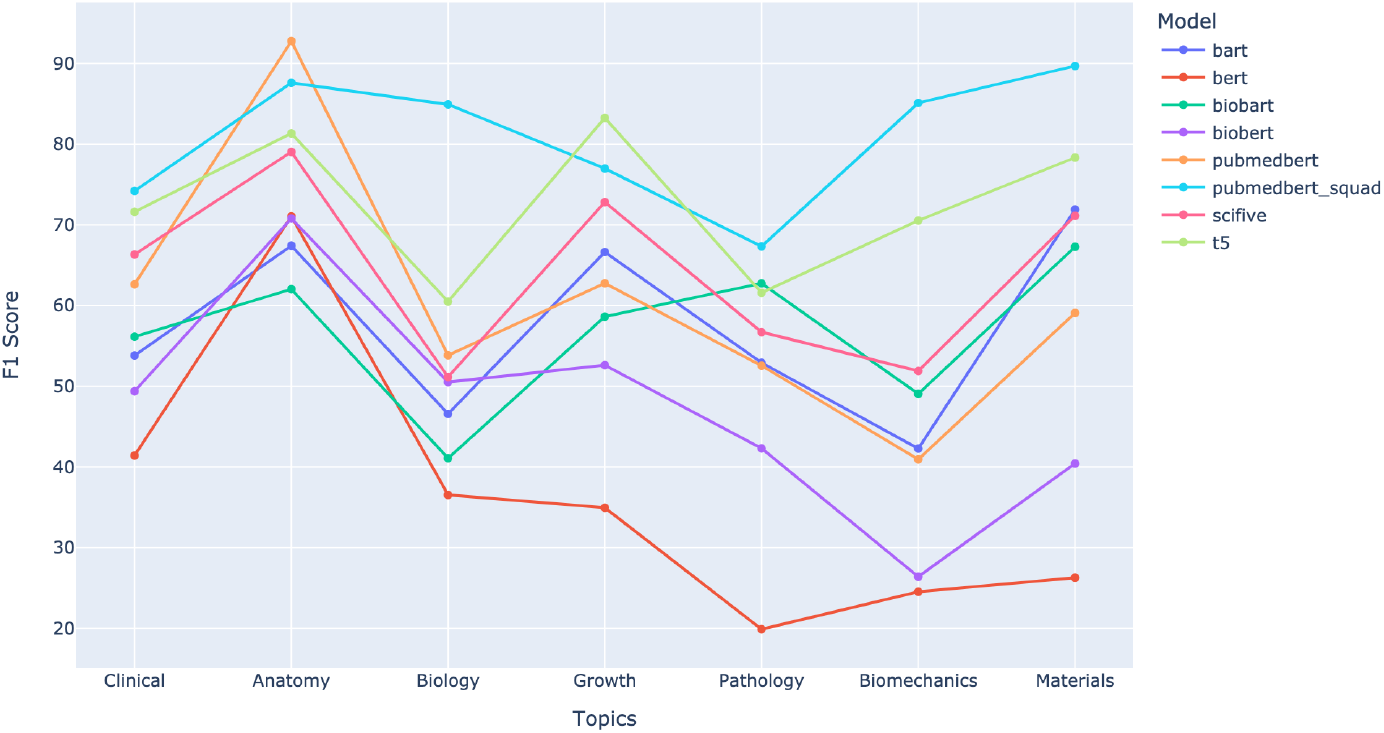
Evaluation by topic of models fine-tuned on OQA. The F1-score of the best model from the three random seed is presented.

## 5 Discussion

### 5.1 Generative models

Contrary to previous reports on few-sample datasets [19], generative models did not yield superior performance to encoder models at a similar parameter count. Performance gains were observed with domain-specific pretraining, with the general domain BERT having the worst performance. When comparing similar models pretrained from scratch, such as PubmedBERT, with a model that underwent continual pretraining, in this case, BioBERT. It was observed that the PubmedBERT model outperformed BioBERT by over 12%. This improvement can be attributed to the use of a vocabulary constructed from PubMed abstracts, which was specifically tailored for biomedical tasks, instead of the vocabulary derived from Wikipedia and BookCorpus used in BioBERT which was generated for the original BERT model. This aligns with the hypothesis that domain-specific vocabulary aids in mastering biomedical tasks [7]. The results from fine-tuning T5 and PubmedBERT_SQuAD_ highlight the importance of leveraging existing large general domain datasets, as these greatly improve transfer learning. In the case of OQA, SQuAD had a very similar format, although no questions pertained to the field of orthodontics. Previous studies reported pre-fine-tuning approaches on SQuAD for biomedical QA [29]. With a similar configuration, the training of PubmedBERT for 2 epochs on this large dataset increased performance by over 16%.

Recent work on teaching LLMs to follow instructions [30, 31] demonstrated improved performance for zero or few-shot transfer. However, these models require considerable computational resources to use. GPT-3.5-turbo (ChatGPT), a closed-source model, belongs to that class. To compare fine-tuned models on specific tasks to an off-the-shelf LLM, zero-shot inference on the OQA test set was performed using the OpenAI API with the “gpt-3.5-turbo” model (August 18th, 2023). The following instructions were provided for each sample: “Answer the following question using the provided context. The answer must correspond to a span of text from the context.”. The resulting F1 score and EM were of 49.77 and 1.75 respectively. It is possible that various prompting techniques could significantly improve performance. However, prompt design can also be technically challenging [32] and unpredictable [33].

### 5.2 Performance variations across models

Averaging the F1 scores for individual test samples across each seed and employing Pearson’s pairwise correlation to compare model performances revealed a general trend of weak correlations between the outputs of most models (Figure 5). An exception was observed among generative models, which showed a notably better correlation within this group. Specifically, the BioBART and SciFive models, following continuous pretraining, exhibited correlations greater than 0.7 when compared to their base models. This pattern may imply that an increase in the scale of pretraining data could potentially lead to some form of convergence in the performance of these models.

**Fig. 5.**
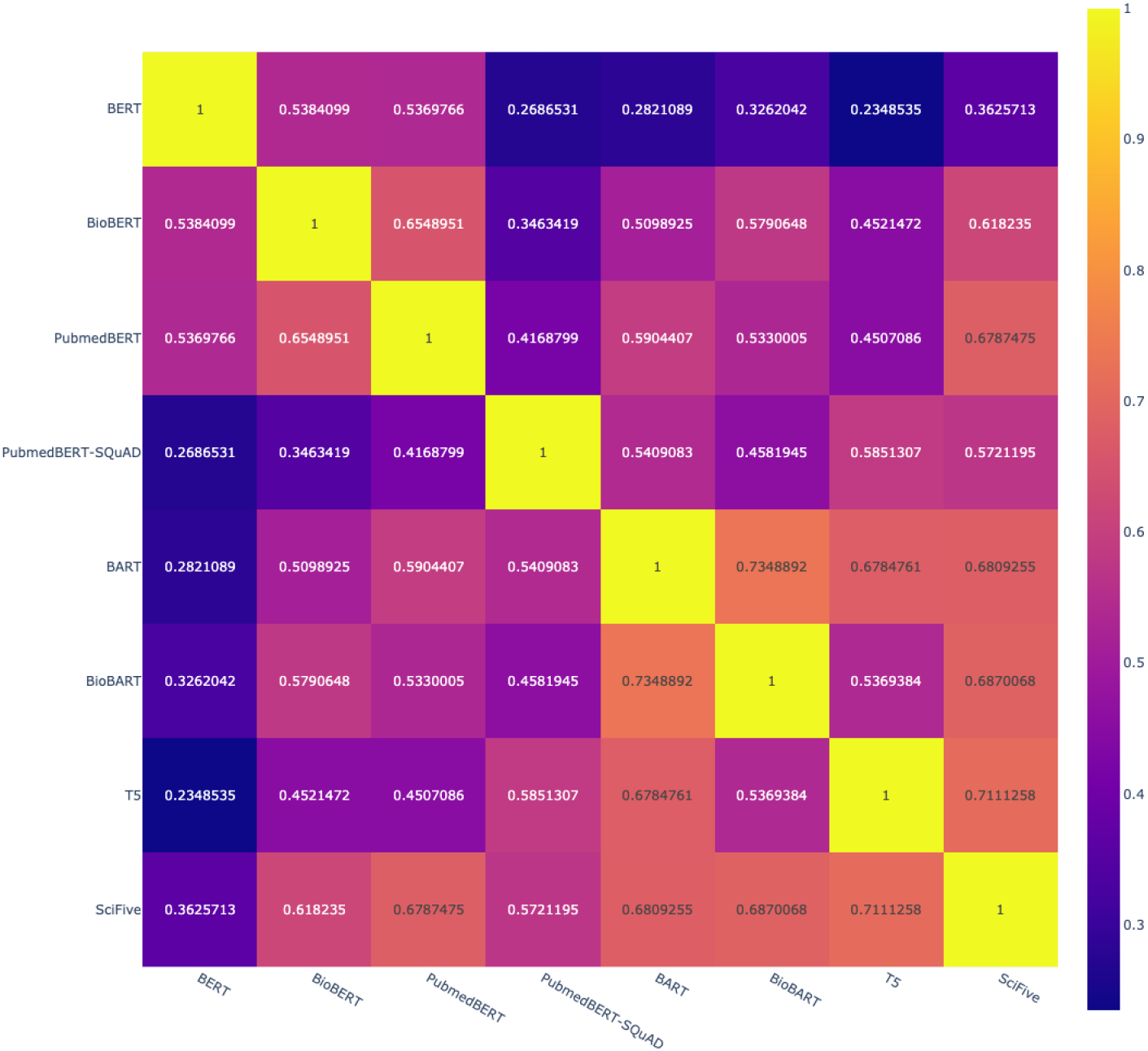
Pairwise correlation of the F1 scores per examples of the test set averaged across seeds (0, 10, 100).

### 5.3 Subgroup analyses

All models performed unevenly across the different topics. Questions related to pathology, biology, and biomechanics were generally less accurate, which respectively accounted for 13.3, 6.47, and 5.28% of the samples in the dataset. Such a drop in performance could be related to their underrepresentation in the dataset. However, the anatomy and materials questions had the highest scores while representing 7.67 and 10.9% of the total number of samples respectively. These variations in performance are not readily explained, as several factors could be at play. Namely, the complexity of the question, LMs lacking background knowledge required to answer the question correctly, or simply the discrepancy between the lengths of the prediction and the label answer. An investigation of performance based on answer length was conducted under the assumption that it could be representative of question complexity. Samples from the test set were grouped into three answer length categories: short (*<*5 tokens), medium (5*<*12 tokens), and long (*>*12 tokens). Although for some models the F1-score varied depending on the answer length, there was not a consistent trend amongst the various fine-tuned models (Figure 6). This phenomenon requires additional investigation incorporating interpretability techniques to diagnose where the issue lies.

**Fig. 6.**
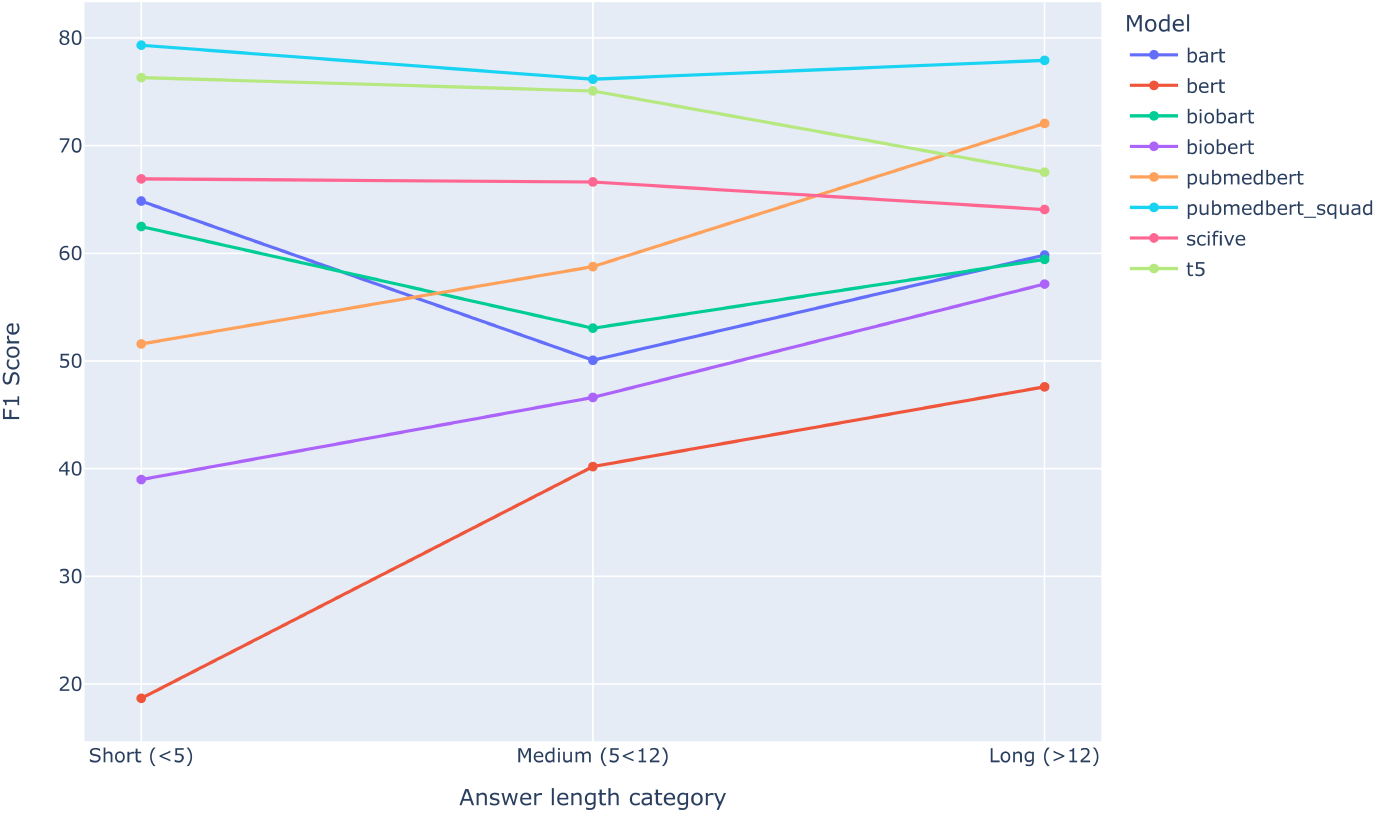
Evaluation by answer length category of models fine-tuned on OQA. The F1-score of the best model from the three random seed is presented.

When assessing the answers qualitatively, the F1 score or EM did not perfectly represent the actual performance of the model in extracting the correct answers. That is, in many cases, the model identified the correct answer but had a poor F1-score or EM simply because the prediction was either too verbose or concise compared to the label answer. Although they generally are a good indicator of model performance, these metrics are not to be mistaken as proxies for “understanding”. A human investigator (MR) evaluated our best model’s output (PubmedBERT_SQuAD_) on all 114 test samples. It was found that 100 predictions were correct and 14 were wrong. Amongst the correct predictions, 15 were verbose and 19 were incomplete when compared to the label answer. This explains why the F1-score was lower than the percentage of questions deemed correct by a human evaluator.

### 5.4 Future work

Recent studies have demonstrated the efficacy of synthetic data in training LMs for general instruction-following tasks [34]. This approach holds promise for expediting the curation process of expert-domain datasets. Automated expert-domain dataset generation techniques represents an interesting area for future research.

## 6 Conclusions

In this article, OrthodonticQA (OQA) the first expert-curated open-access QA dataset specific to orthodontics was presented. A general template-based approach to question formulation incorporating PICO information was presented. A diverse selection of state-of-the-art language models were fine-tuned and evaluated performance on OQA. Post-hoc analyses highlight considerable variations in performance depending on topic subgroups, the importance of pre-fine-tuning on large QA datasets and the need for human evaluation of outputs to correctly assess capabilities.

## Data Availability

All data produced are available online at https://huggingface.co/datasets/m-rousseau/oqa-v1

https://github.com/maxrousseau/o-nlp

## Declarations

### Ethics approval and consent to participate

Not applicable.

## Consent for publication

Not applicable.

## Availability of data and materials

Both the OQA dataset and code to reproduce our experiments are made available under the open-source Apache-2.0 license.

- source code repository: https://github.com/maxrousseau/o-nlp
- dataset repository: https://huggingface.co/datasets/m-rousseau/oqa-v1

## Acknowledgments

The main author would like to acknowledge the financial support provided by the University of Montreal Faculty of Dentistry to conduct this research. This research was enabled in part by support provided by Calcul Québec (https://www.calculquebec.ca/) and the Digital Research Alliance of Canada (alliancecan.ca).

## Footnotes

1 https://huggingface.co/datasets/m-rousseau/oqa-v1

2 https://meridian.allenpress.com/angle-orthodontist

3 (https://github.com/maxrousseau/o-nlp)

## Notes

### Competing Interest Statement

The authors have declared no competing interest.

### Summary of Updates

Fixed typo in author affiliations.

